# *Identification and Characterization of Rare Pathogenic* Gene*tic Variants Associated with Breast Cancer Susceptibility in 1000 Genome and gnomAD Population*

**DOI:** 10.1101/2025.08.30.25334747

**Authors:** Kanishk Yadav, Abhishek Prasad

## Abstract

**Background:** Inherited variants in cancer susceptibility genes play a critical role in breast cancer risk, yet comprehensive comparative analyses across clinically relevant genes remain limited. While *BRCA1* and *BRCA2* are well-established high-risk contributors, the broader landscape of germline pathogenicity across breast cancer associated genes warrants systematic evaluation.

**Methods:** We curated a panel of 15 breast cancer associated genes (*BRCA1*, *BRCA2*, *TP53*, *PTEN*, *CHEK2*, *ATM*, *CDH1*, *PALB2*, *LSP1*, *MAP3K1*, *NF1*, *RECQL*, *TOX3*, *FANCD2*, *RAD51C*) based on clinical and epidemiological relevance. Publicly available germline variant data from gnomAD v4.1 and the 1000 Genomes Project were integrated with ClinVar annotations. Variants were classified by clinical significance and filtered using allele frequency and phenotype annotation to stratify into high-risk and non-high-risk groups. We compared the distribution of variants and assessed penetrance patterns, and we evaluated the discriminatory power of computational pathogenicity predictors including CADD, REVEL, SIFT, and PolyPhen-2.

**Results:** *BRCA1* and *BRCA2* harbored the highest burden of pathogenic variants, consistent with their high penetrance status. Moderate-risk genes (*PALB2*, *RAD51C*, *CHEK2*) showed intermediate levels of pathogenicity, while low-risk genes such as *RECQL*, *TOX3*, and *FANCD2* were predominantly enriched for variants of uncertain significance. High-risk variants exhibited significantly elevated CADD (>30), REVEL (>0.75), and SIFT (<0.05) scores compared to non-high-risk variants (p < 1×10^2^), whereas PolyPhen-2 showed limited discriminatory power. Odds ratio analysis further supported CADD, REVEL, and ClinVar pathogenicity labels as strong discriminators between variant categories.

**Conclusions:** Our analysis reaffirms the pathogenic burden concentrated in high penetrance genes and highlights the utility of combined in silico prediction tools and population frequency filters in variant prioritization. These findings support refined variant curation pipelines for hereditary breast cancer and provide a framework for interpreting gene-level pathogenicity across diverse susceptibility loci.

## Introduction

Breast cancer is the most frequently diagnosed cancer in women worldwide and a leading cause of female cancer related mortality. In 2020, there were approximately 2.3 million new cases and over 670,000 deaths attributed to breast cancer globally, illustrating the significant burden of this disease [1]. Although the majority of breast cancers are sporadic, 5–10% are hereditary, and an even larger proportion may involve inherited susceptibility in familial clusters. In certain populations, up to 15–25% of breast cancer cases may be associated with germline mutations in cancer predisposition genes [2,27]. Historically, *BRCA1* and *BRCA2* have been the most studied genes, with pathogenic variants conferring lifetime breast cancer risks as high as 65% for *BRCA1* and 57% for *BRCA2* [6,7]. Other high penetrance genes, such as *TP53*, *PTEN*, and *CDH1*, are associated with hereditary cancer syndromes like Li-Fraumeni, Cowden, and hereditary diffuse gastric cancer [8–10].

Beyond these, moderate risk genes such as *PALB2*, *CHEK2*, *ATM*, and *RAD51C* also play clinically significant roles in genetic testing panels and contribute additional, though smaller, risk burdens [11–14]. The ClinVar database has become an essential open access resource for clinical variant interpretation, consolidating expert submissions, laboratory assertions, and supporting evidence [1,2]. However, interpretation challenges persist, variants of uncertain significance (VUS) still make up a large proportion of classified variants in cancer susceptibility genes [1,5,9]. Although ClinGen continues to refine gene and variant classification frameworks through its Variant and Gene Curation Expert Panels [23,41], no single universally accepted breast cancer gene panel exists for research standardization. Instead, researchers rely on curated sources such as the National Cancer Institute’s PDQ Genetics summary and systematic reviews of genetic contributions to breast cancer to define working gene sets [27,28].

In this study, we leveraged population scale germline variant resources from gnomAD v4.1 and the 1000 Genomes Project [24,29] to extract and annotate variants across 15 breast cancer associated genes frequently referenced in clinical and research settings: *BRCA1*, *BRCA2*, *TP53*, *PTEN*, *CHEK2*, *ATM*, *CDH1*, *PALB2*, *LSP1*, *MAP3K1*, *NF1*, *RECQL*, *TOX3*, *FANCD2*, and *RAD51C* [2,27,28]. We then harmonized this data with ClinVar to evaluate the distribution of clinical significance classifications, stratified variants into high risk and non high risk categories based on frequency and annotation thresholds, and compared the effectiveness of in silico pathogenicity predictors such as CADD, REVEL, SIFT, and PolyPhen-2 in distinguishing variant classes [18–22].

## Materials and Methods

### Data sources and gene set

We aggregated germline single-nucleotide variants (SNVs) for breast-cancer associated genes from public resources: 1000 Genomes Project and gnomAD v4 (GRCh38) (accessed Aug 2025) [24,29]. Gene selection was guided by a recent peer-reviewed review and the NCI PDQ professional guideline, yielding the following set: *BRCA1*, *BRCA2*, *TP53*, *PTEN*, *CHEK2*, *ATM*, *CDH1*, *PALB2*, *LSP1*, *MAP3K1*, *NF1*, *RECQL*, *TOX3*, *FANCD2*, *RAD51C* [27,28]. Gene symbols followed HGNC standards to ensure consistent nomenclature across datasets [30]. To reduce potential errors, we verified gene lists against Ensembl BioMart and RefSeq identifiers, ensuring that gene boundaries and canonical transcripts were consistent with current reference annotations. This cross-check minimized misassignment of variants to paralogous loci, which can occur in large-scale databases.(Flowchart 1)

**Flowchart 1.**
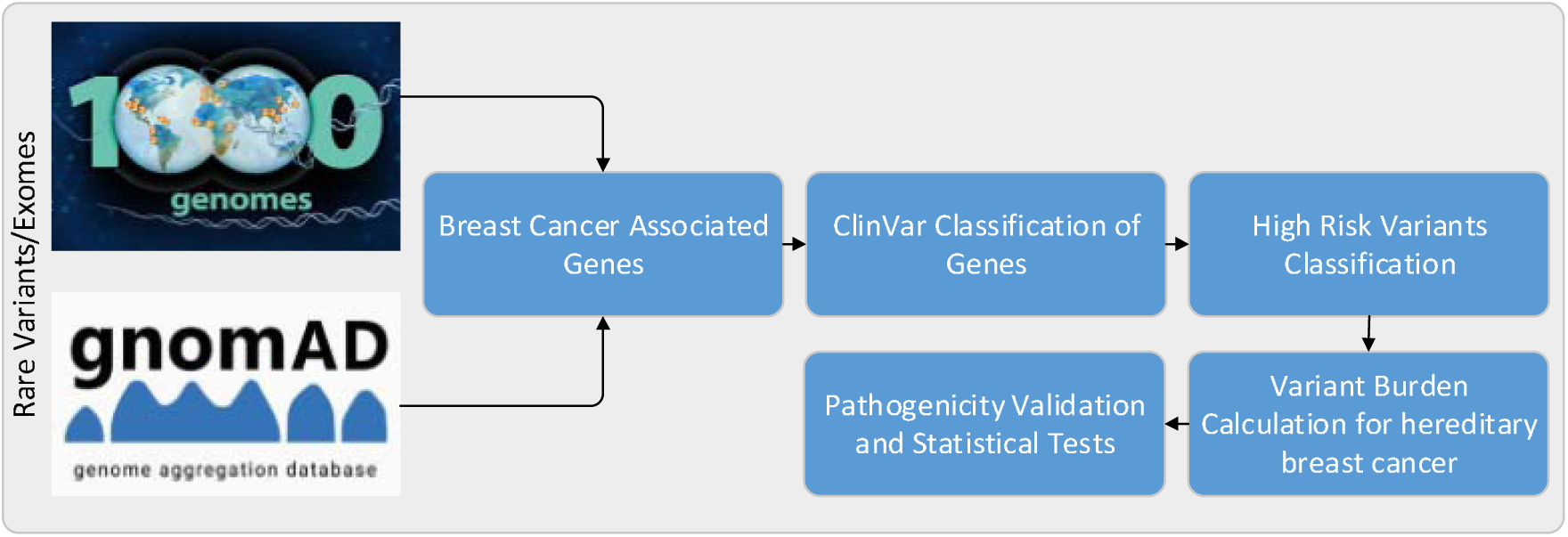
Overview of the analytical pipeline used to identify and evaluate high-risk germline variants associated with hereditary breast cancer. Variant data from the 1000 Genomes Project and gnomAD v4 were used to extract rare single-nucleotide variants (SNVs) within a curated set of breast cancer susceptibility genes. These variants were annotated using ClinVar to classify their clinical significance, followed by identification of high-risk variants based on allele frequency, disease relevance, and predicted pathogenicity. Subsequent statistical validation and burden analyses were performed to quantify variant impact across genes.

### Variant harmonization and ClinVar Labeling

All variants were left-joined to ClinVar to obtain clinical significance (CLNSIG) labels (Pathogenic [P], Likely Pathogenic [LP], P/LP, VUS, Likely Benign [LB], Benign/Likely Benign [B/LB], Benign [B]) using dbSNP rsIDs (avsnp151) as the stable key [1,2,26]. Variants lacking an rsID (avsnp151 = “.”) were excluded from Figure 1 to maintain uniqueness and cross-database comparability [26]. When the same rsID/gene appeared with discordant CLNSIG labels (e.g., both P and LP), we resolved to a single most-specific/most-severe label using the deterministic priority P > LP > P/LP > VUS > LB > B/LB > B (implemented by sorting on a numeric priority and retaining the first record). This rule privileges definitive calls (P) over ambiguous or benign classifications and avoids double-counting. Additional harmonization included collapsing redundant ClinVar submissions under the same rsID, thereby reducing inflation from multiple submitter entries.

**Figure 1.**
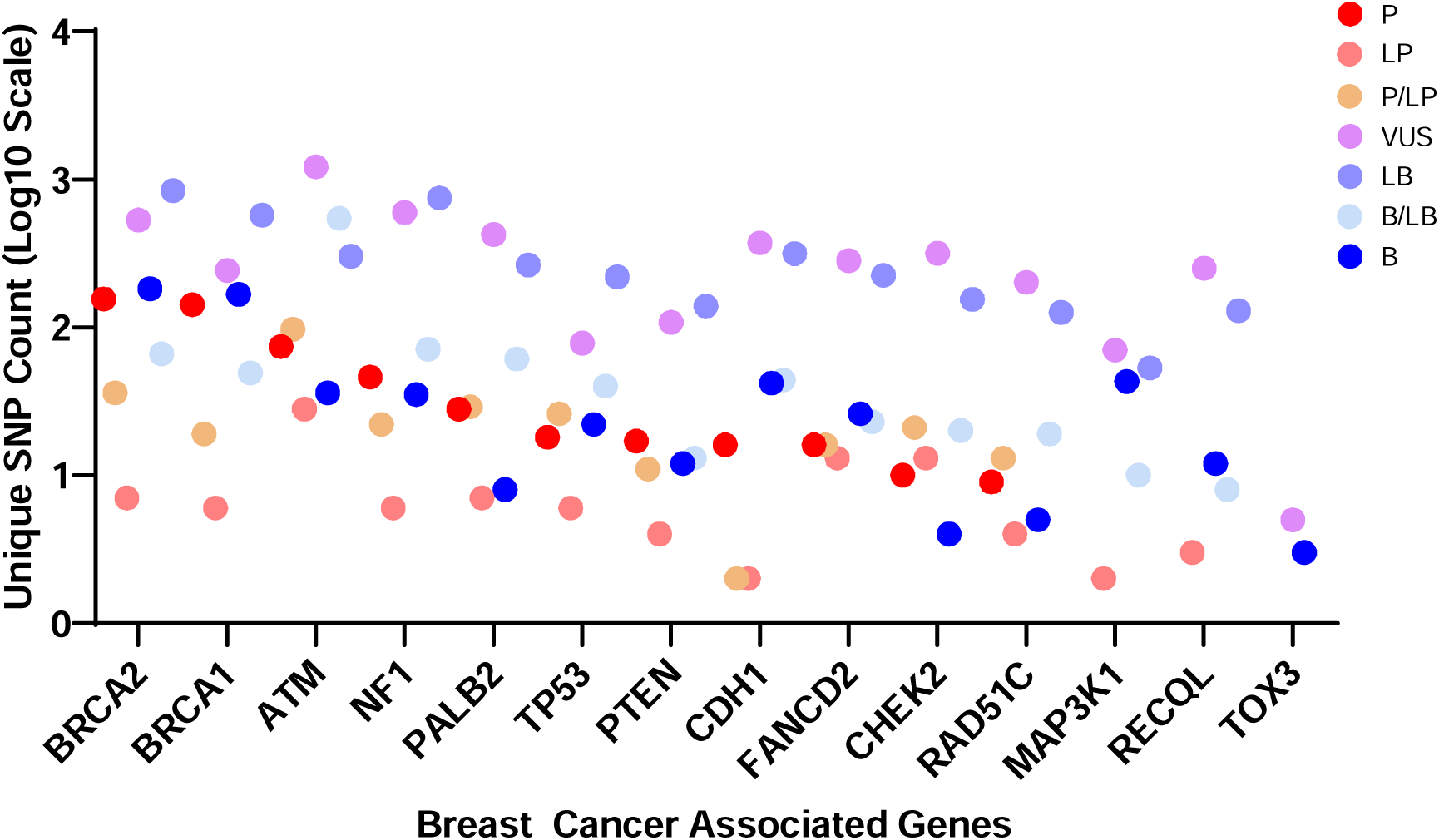
Distribution of ClinVar-classified variants across breast cancer susceptibility genes. Log-scaled counts of unique SNPs per gene are shown, colored by clinical significance (P, LP, P/LP, VUS, LB, B/LB, B). *BRCA1* and *BRCA2* show the highest pathogenic variant counts, while low-penetrance genes like *MAP3K1*, *RECQL*, and *TOX3* have few. VUS remain prevalent across genes, reflecting challenges in interpretation.

### Counting unique variants per gene and CLNSIG class

After harmonization and de-duplication by (rsID, gene), we computed unique rsID counts per gene within each CLNSIG class. Counts reflect the number of distinct dbSNP identifiers in a given gene/CLNSIG bin, not alleles or submissions. This yields a gene-by-class matrix used to visualize the distribution of reported variation across 15 gene*s* and 7 ClinVar categories. To ensure robustness, we also verified that the same variant did not map to multiple gene*s* due to overlapping annotations, and any such ambiguous cases were excluded. The resulting dataset captures the breadth of known germline variation across clinically relevant breast cancer gene*s*, emphasizing pathogenic and likely pathogenic variants while contextualizing them within the full spectrum of ClinVar labels.

### Defining the high-risk variants with heavy variant burden

We defined the high-risk candidate set ‘Cases’ that simultaneously met three criteria: (i) rarity, operationalized as gnomAD exome allele frequency ≤1% [24,25]; (ii) disease relevance, requiring the ClinVar Diagnosis term (CLNDN) to contain “Hereditary_breast” (e.g., “Hereditary_breast_and_ovarian_cancer”) [1]; and (iii) predicted deleteriousness, with CADD PHRED ≥30 to enrich for highly damaging changes [18,19]. (Thresholds for other predictors such as REVEL were not used to define Cases for Figure 2 and were evaluated separately in Figure 3.) To quantify burden, we first deduplicated records at the (rsID, gene) level, because a single variant can have multiple submissions/labels—by retaining one clinical label using a deterministic severity priority: P > P/LP > LP > VUS > LB > B/LB > B [1,2]. We then constructed, for each (gene, CLNSIG) bin, a numerator equal to the count of unique rsIDs among Cases (***N***_{cases}_) and a denominator equal to the count of all unique rsIDs after deduplication (***N***_{total}_). The “variant risk burden” was computed as 100 × (numerator/denominator); missing cells were imputed as 0. This measure reflects the proportion of catalogued variants within each gene/class that satisfy our rare, disease-linked, high-deleteriousness filter and is not a clinical penetrance estimate.

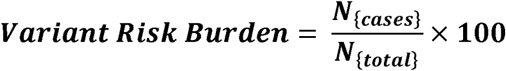

**Figure 2.**
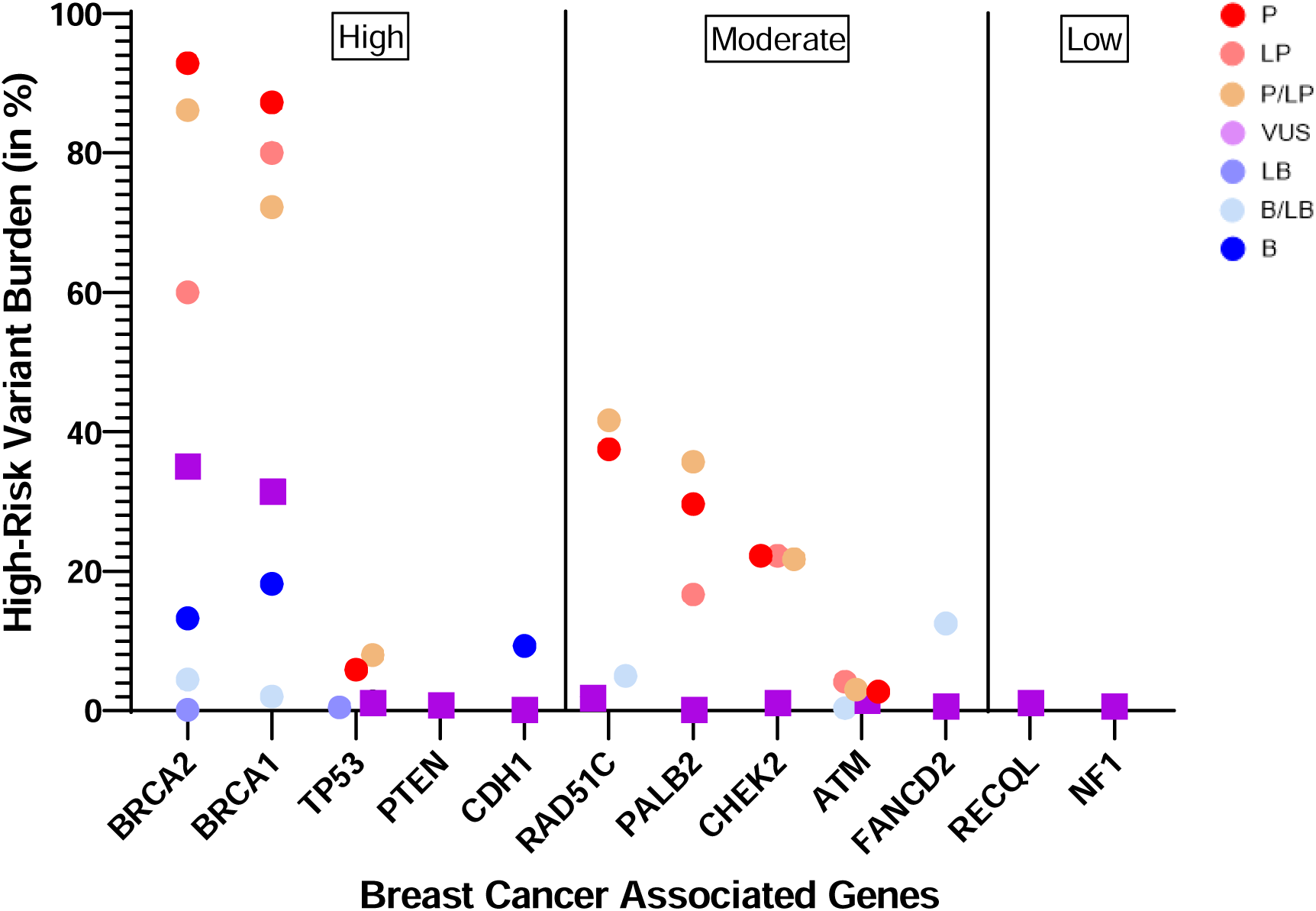
*High-risk variant burden across breast cancer susceptibility* gene*s.* Percentage of high-risk variants (Pathogenic, Likely Pathogenic, and P/LP) per gene, stratified by penetrance class (high, moderate, low). Genes like *BRCA2* and *BRCA1* showed the highest burden (>85%), followed by *TP53*. Moderate-penetrance genes such as *PALB2* and *RAD51C* had intermediate burdens (∼30–40%), while low-penetrance genes (*NF1*, *RECQL*, *FANCD2*) had minimal high-risk burden and were dominated by VUS.

**Figure 3.**
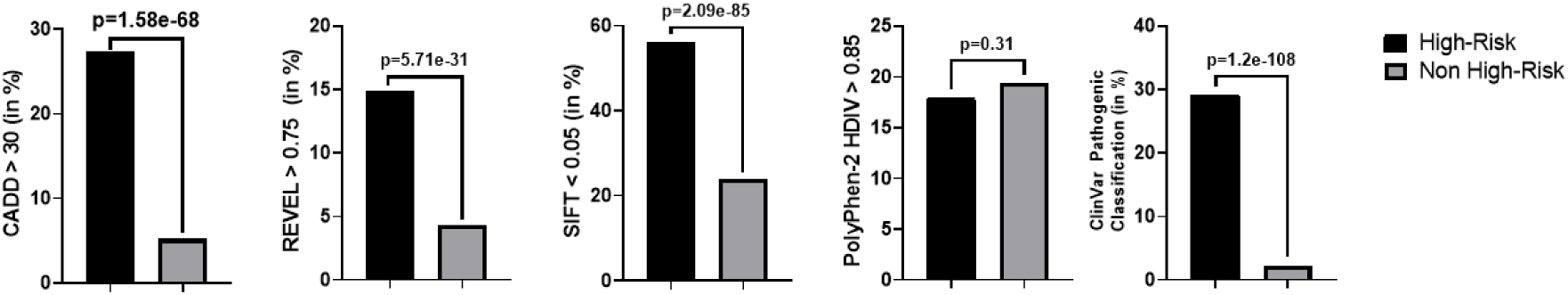
Comparison of in silico pathogenicity scores between high-risk and non-high-risk variants. High-risk variants showed significantly higher proportions exceeding established cutoffs for CADD (>30), REVEL (>0.75), and SIFT (<0.05) compared to non-high-risk variants (all p<10^3^). No significant difference was observed for PolyPhen-2 HDIV (>0.85). ClinVar pathogenic classifications were also markedly enriched among high-risk variants (33.8% vs 1.9%, p=1.2×10^1^). These findings support the utility of select in silico scores and ClinVar assertions in distinguishing clinically relevant variants.

Genes were assigned to high, moderate, or low penetrance classes using prior risk estimates [3–5]. For display, genes were ordered within each class by descending Pathogenic (P) burden, then VUS, then Benign (B) to emphasize clinically actionable content. We produced a stacked bar plot showing B, VUS, and P percentages per gene with dashed separators and labels for the three penetrance strata. Axes were labeled “Breast Cancer Associated Genes” (x) and “Variant Risk Burden (%)” (y) (Figure 2). To improve interpretability, we further highlighted the most frequently mutated genes within each penetrance category, providing a visual emphasis on loci of particular clinical and biological relevance. The choice of stacked percentages (rather than absolute counts) allows for fairer comparison across genes with different total variant loads. Color schemes were selected to align with conventional clinical classification (e.g., red for pathogenic and moving down to blue for benign), thereby facilitating intuitive interpretation for both research and clinical audiences.

### Rationale and QC

Using AF ≤ 1% focuses on rare alleles that are more likely pathogenic in Mendelian cancer genes [24,25]. A stringent CADD ≥ 30 enriches for variants with strong predicted functional impact [18,19]. Severity-based deduplication using dbSNP rsIDs ensures one vote per variant per gene and reduces bias from multiple submissions [1,2,26]. In addition, we excluded synonymous and intronic variants outside canonical splice sites to reduce noise from variants unlikely to alter protein function. QC steps also included verification of gene-variant mappings against multiple annotation sources (ANNOVAR, ClinVar, and gnomAD) to minimize misclassification errors. Where multiple isoforms existed, we prioritized canonical transcripts as defined by Ensembl for consistency across genes. Variant counts were cross-checked by independent scripts to ensure reproducibility of the burden profiles presented in Figure 2. All steps were performed in Python with pandas/NumPy for reproducibility [31–33]. Computational notebooks and code were version-controlled, with random seeds fixed where applicable, ensuring complete reproducibility and transparency of analyses.

### Validation of the pathogenicity contributing factors in the for high-risk vs. non high-risk burden variants

To compare computational pathogenicity signal between high-risk and non-high-risk variant groups, we partitioned the severity-prioritized, de-duplicated rsID–gene set and, for each group, computed the proportion of unique variants exceeding established cut-points for widely used predictors—CADD PHRED >30, REVEL >0.75, SIFT <0.05 (damaging), PolyPhen-2 HDIV >0.85 (probably damaging)—and the proportion annotated Pathogenic in ClinVar. Proportions were calculated as 100 × {n=unique rsIDs meeting threshold}/{N=unique rsIDs in group} across all 15 genes. Between-group differences were assessed with two-sided Fisher’s exact tests, and effect sizes were summarized as odds ratios (ORs) with 95% CIs using log(OR) (Woolf) approximations (Figure 4). Thresholds reflect original tool validations, systematic cross-tool evaluations, and contemporary ClinGen SVI guidance/calibration on the use and evidentiary weight of in-silico predictors (notably PP3/BP4), as well as dbNSFP aggregator releases that standardize predictor score retrieval and recommended cut-points [1,2,18–22,34–41,42,43]. Key benchmarking and guidance sources include Genome Biology’s systematic review of in-silico algorithm use under ACMG/AMP [34], studies highlighting evaluation pitfalls and circularity [35], comparative and meta-predictor analyses [36,38], the dbNSFP v4 compendium [37], cancer-gene functional assay benchmarking with likelihood ratios for 44 tools [39], ClinGen SVI calibration of PP3/BP4 and recommendations for single pre-specified tools [40], and a Bayesian formalization of ACMG/AMP combining rules [41], with recent gene-by-gene calibration of REVEL/BayesDel further motivating fixed a priori thresholds [9].

**Figure 4.**
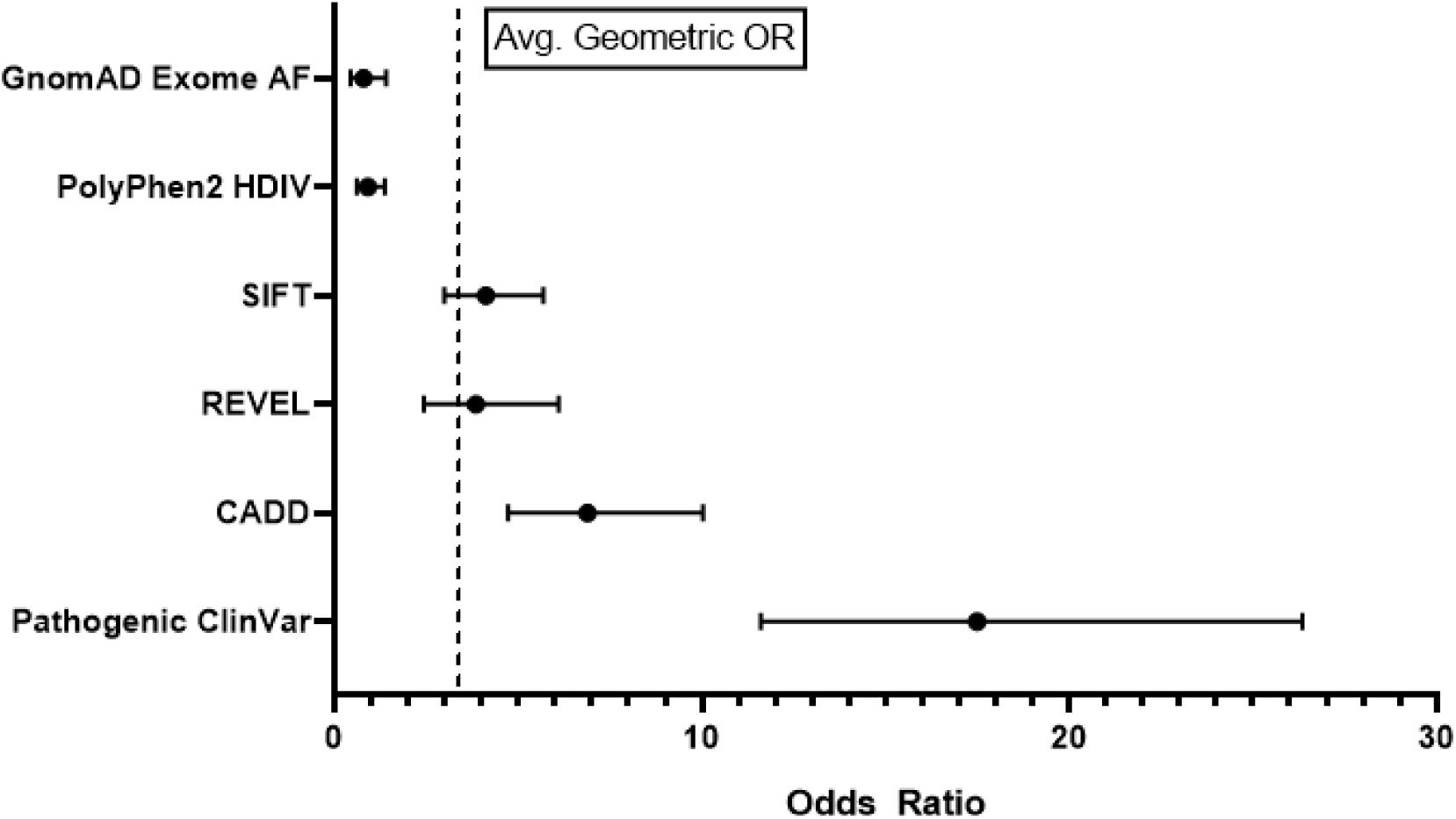
Odds ratios (ORs) with 95% confidence intervals (CIs) for high-risk versus non-high-risk variants across key predictors. The ClinVar pathogenic classification demonstrated the strongest discriminatory association (geometric mean OR ≈ 17.5), followed by CADD (≈ 6.9), SIFT (≈ 4.2), and REVEL (≈ 3.8). In contrast, PolyPhen-2 HDIV (≈ 0.9) and gnomAD exome allele frequency (≈ 0.8) were weak or non-informative in distinguishing high-risk variants. ORs were estimated using 2×2 Fisher’s exact tests; point estimates represent geometric means across three replicates, and error bars denote observed minimum and maximum ORs.

## Results

### Distribution of ClinVar-Classified Variants across Breast Cancer Susceptibility Genes

We evaluated the unique ClinVar-reported SNPs across 15 breast cancer susceptibility genes, categorized by ClinVar clinical significance (Figure 1). Variant counts varied widely between genes, with *BRCA2* and *BRCA1* showing the highest numbers (>100 unique SNPs), followed by *ATM, NF1, PALB2, and TP53*, whereas *MAP3K1*, *RECQL*, *and TOX3* had very few unique SNPs. The proportion of entries classified as VUS remains substantial in ClinVar for cancer-associated genes, reflecting ongoing challenges in clinical interpretation [1,2]. Figure 1 demonstrates a declining gradient in total variant counts and in the proportion of pathogenic variants from high-penetrance genes (e.g*., BRCA2 to PALB2 to TP53*) to moderate (*CHEK2*, *RAD51C*) and low-penetrance loci (*MAP3K1, RECQL, TOX3*). These patterns are consistent with large studies showing that pathogenic variants are identified in a meaningful fraction of breast-cancer cases, with *BRCA1/2* contributing the largest share and additional risk arising from other susceptibility genes [3–5]. Figure 1 clearly demonstrates a declining gradient in both total variant counts and the proportion of pathogenic variants, from high-penetrance (e.g., *BRCA2 to PALB2 to TP53*) to moderate (*CHEK2, RAD51C*) and low-penetrance loci (*MAP3K1, RECQL, TOX3*).

We next quantified the burden of high-risk variants for hereditary breast cancer as defined in the Methods (Pathogenic [P], Likely Pathogenic [LP], and combined P/LP) across the 15 genes (Figure 2; Table 1). Genes were categorized into high, moderate, and low penetrance groups based on established risk estimates from prior studies [3–5]. High-penetrance genes showed the greatest burden of pathogenic variants. *BRCA2* exhibited the highest proportion of P variants (92.86%), followed by *BRCA1* (87.23%) and *TP53* (5.88%) [6–8]. Similarly, P/LP combined frequencies were highest in *BRCA2* (86.11%) and *BRCA1* (72.22%), with *TP53* markedly lower (8.00%) [6–8]. *PTEN* and *CDH1* also carried clinically relevant high-risk variants, albeit at lower frequencies than *BRCA1/2* [9,10]. In the moderate-penetrance category, *PALB2* and *RAD51C* had the highest pathogenic variant proportions (29.63% and 37.00%, respectively), followed by *CHEK2* (22.22%) [11–13]. *ATM* carried a comparatively low fraction of pathogenic variants (2.74%) [14]. While *PALB2* and *RAD51C* showed substantial P/LP burdens (>35%), the remaining moderate-penetrance genes exhibited lower percentages [11–14]. For low-penetrance genes, the proportion of pathogenic variants was negligible, with most loci—such as *NF1*, *RECQL*, and Fanconi-pathway genes—showing ≤1–2% high-risk variant burden; most variants were VUS [15–17]. Overall, these analyses highlight a gradient in pathogenic burden that mirrors penetrance categories and agree with large panel and population studies reporting that *BRCA1/2* account for most actionable hereditary breast-cancer variants, with additional but smaller contributions from non-BRCA genes [3–5].In Silico Pathogenicity Predictions for High-Risk versus Non-High-Risk Variants

**Table 1.** Variant Burden of genes by their ClinVar classification for determined High-Risk variants threshold for hereditary breast cancer:

| Gene | B | B/LB | LB | LP | P | P/LP | VUS |
| --- | --- | --- | --- | --- | --- | --- | --- |
| *ATM* | 0 | 0.362319 | 0 | 4.166667 | 2.739726 | 3.030303 | 1.486375 |
| *BRCA1* | 18.18182 | 2.040816 | 0 | 80 | 87.23404 | 72.22222 | 31.53527 |
| *BRCA2* | 13.26531 | 4.477612 | 0.1221 | 60 | 92.85714 | 86.11111 | 35.16068 |
| *CDH1* | 9.302326 | 0 | 0 | 0 | 0 | 0 | 0.27027 |
| *CHEK2* | 0 | 0 | 0 | 22.22222 | 22.22222 | 21.73913 | 1.269841 |
| *FANCD2* | 0 | 12.5 | 0 | 0 | 0 | 0 | 0.714286 |
| *MAP3K1* | 0 | 0 | 0 | 0 | 0 | 0 | 0 |
| *NF1* | 0 | 0 | 0 | 0 | 0 | 0 | 0.671141 |
| *PALB2* | 0 | 0 | 0 | 16.66667 | 29.62963 | 35.71429 | 0.236407 |
| *PTEN* | 0 | 0 | 0 | 0 | 0 | 0 | 0.934579 |
| *RAD51C* | 0 | 5 | 0 | 0 | 37.5 | 41.66667 | 2 |
| *RECQL* | 0 | 0 | 0 | 0 | 0 | 0 | 1.204819 |
| *TOX3* | 0 | 0 | 0 | 0 | 0 | 0 | 0 |
| *TP53* | 0 | 0 | 0.47619 | 0 | 5.882353 | 8 | 1.298701 |

### *In-Silico* Pathogenicity Predictions (High-Risk vs Non-High-Risk)

We compared multiple in silico scores between high-risk and non-high-risk variants (Figure 3). High-risk variants had a greater proportion with CADD > 30 (27.9% vs 6.5%, p = 1.58×10) [18,19] and REVEL > 0.75 (15.4% vs 4.1%, p = 5.71×10^31^) [20]. A significantly larger fraction of high-risk variants had SIFT < 0.05 (57.3% vs 23.6%, p = 2.09×10) [21]. PolyPhen-2 HDIV > 0.85 was not different between groups (18.4% vs 19.7%, p = 0.31) [22]. As expected, the proportion of variants classified as pathogenic in ClinVar was far higher in the high-risk group (33.8% vs 1.9%, p = 1.20×10^1^) [1,23]. Together, CADD, REVEL, and SIFT showed stronger discrimination than PolyPhen-2 HDIV, with ClinVar classifications aligning with our high-risk grouping.

### Odds Ratios for Pathogenicity Predictors in High-Risk versus Non-High-Risk Variants

To quantify discriminatory performance between high-risk and non-high-risk variant groups, we estimated odds ratios (ORs) with 95% confidence intervals (CIs) for each predictor using two-sided Fisher’s exact tests (Figure 4). ClinVar pathogenic classification yielded the strongest association, with ORs ranging from 14.81 to 20.60 and a geometric mean of approximately 17.54, highlighting its value as a clinical gold standard for identifying pathogenic variants [1,23]. Among computational prediction tools, CADD showed the highest discriminatory power (geometric mean OR ≈ 6.92), reflecting its integration of diverse annotation, functional, and conservation-based features into a genome-wide pathogenicity score [18,19]. SIFT and REVEL followed with moderate associations (geometric means ≈ 4.23 and 3.84, respectively), indicating that protein conservation and ensemble-based models are informative in predicting variant impact [20,21]. In contrast, PolyPhen-2 HDIV exhibited limited discriminatory capacity (geometric mean OR ≈ 0.90), consistent with prior evaluations reporting lower performance and calibration for clinical use [22,34–36].

gnomAD exome allele frequency yielded a geometric mean OR of 0.80, indicating that although high-risk variants are typically rare, rarity alone is insufficient to distinguish pathogenicity without supporting clinical or functional annotation [24,25]. Overall, these results emphasize the importance of integrating multiple lines of evidence for variant classification and suggest that while some prediction tools are informative, none alone can substitute for curated clinical databases or experimentally validated evidence.

## Discussion

This study presents a comprehensive computational analysis of germline single-nucleotide variants (SNVs) in 15 breast cancer susceptibility genes using publicly available variant databases and ClinVar annotations. By integrating pathogenicity predictors (e.g., CADD, REVEL, SIFT, PolyPhen-2 HDIV) with allele frequency data from gnomAD and disease associations from ClinVar, we quantified the burden and distribution of high-risk variants across genes and assessed the discriminatory power of commonly used in silico tools.

Our results reaffirm the dominant contribution of *BRCA1* and *BRCA2* to hereditary breast cancer risk, with both genes showing the highest number of unique pathogenic or likely pathogenic (P/LP) variants and the greatest high-risk variant burden. This aligns with decades of literature identifying *BRCA1/2* mutations as the most penetrant drivers of familial breast cancer, with lifetime risk estimates ranging from 45% to 70% [6,7]. Genes such as *TP53*, *PTEN*, and *CDH1* also harbored clinically relevant high-risk variants but at lower frequencies, consistent with their established classification as high-penetrance genes involved in syndromic cancer risk (e.g., Li-Fraumeni and Cowden syndromes) [8–10]. Among moderate-penetrance genes, *PALB2*, *CHEK2*, and *RAD51C* showed measurable pathogenic variant frequencies, in agreement with population-based sequencing studies [11–13].

The observed gradient in pathogenic variant burden, from high to moderate to low-penetrance genes—is concordant with prior risk modeling studies [3–5]. For example, *ATM* and *FANCD2* showed low P/LP proportions, while *NF1*, *RECQL*, and *TOX3* had negligible high-risk variant content, suggesting that their overall contribution to inherited breast cancer may be limited or context-dependent [14–17]. Notably, *NF1* has previously been associated with moderate risk in specific subpopulations, though the majority of variants remain of uncertain significance [15]. When stratifying variants into high-risk versus non-high-risk groups based on allele frequency, disease label, and deleteriousness (CADD ≥ 30), we observed significant enrichment of functional impact signals in the high-risk group. CADD, REVEL, and SIFT scores were significantly elevated among high-risk variants, with odds ratios ranging from 3.8 to 6.9, while PolyPhen-2 HDIV showed no appreciable difference. These findings are in line with prior evaluations of pathogenicity prediction tools, which have consistently identified CADD and REVEL as among the best-performing algorithms for rare variant classification [34–38]. Our results also reinforce the limitations of relying on a single in silico tool and underscore the importance of integrating multiple evidence lines, as outlined in ACMG/AMP variant interpretation guidelines [23,40].

ClinVar pathogenic classification emerged as the most powerful discriminator of high-risk status, with an odds ratio exceeding 17 and a highly significant p-value (p < 1e-100). This reflects the clinical utility of curated variant databases and expert-reviewed annotations.

However, our findings also highlight persistent challenges: the high proportion of VUS across many genes reflects the uncertainty still surrounding rare missense variants and the need for more functional data, particularly in under-characterized loci. Despite the strengths of this analysis, including rigorous variant deduplication and prioritization pipelines, there are limitations. First, ClinVar submissions may be biased toward well-studied genes such as *BRCA1/2*, potentially inflating their apparent variant burden. Second, pathogenicity predictors are trained largely on known variants and may be less accurate for genes with sparse functional data. Third, the reliance on public databases limits clinical granularity (e.g., family history, co-occurring mutations, ancestry), which are critical for accurate risk modeling.

Future directions include incorporation of larger-scale case-control sequencing datasets, functional assay data, and population-specific variant annotations. As variant databases continue to grow, harmonized frameworks combining clinical data, evolutionary conservation, functional assays, and machine learning approaches will be key to resolving the large pool of VUS and refining gene-level penetrance estimates.

In conclusion, our study reinforces the centrality of *BRCA1/2* in hereditary breast cancer risk while providing comparative insight into other susceptibility genes. Computational pathogenicity scores such as CADD and REVEL effectively enrich for high-risk variants and may serve as supportive evidence in variant interpretation. However, their performance varies, and integrating them with curated clinical databases remains essential for robust classification. These findings support ongoing efforts to calibrate in silico tools and advocate for standardized, evidence-based variant assessment pipelines in clinical genomics.

## Conclusion

This study presents a comprehensive variant-level analysis of 15 breast cancer susceptibility genes using large public databases and ClinVar annotations. Our findings reaffirm the central role of *BRCA1* and *BRCA2*, which exhibited the highest burden of high-risk variants towards hereditary breast cancer, consistent with their well-established high penetrance. Moderate-penetrance genes such as *PALB2*, *CHEK2*, and *RAD51C* showed appreciable pathogenic variant burdens, supporting their inclusion in clinical testing panels. In contrast, low-penetrance genes like *MAP3K1*, *RECQL*, and *TOX3* were predominantly enriched for variants of uncertain significance, highlighting gaps in current variant interpretation efforts.

In silico tools such as CADD, REVEL, and SIFT effectively discriminated high-risk from non-high-risk variants, with significantly higher proportions of high-scoring variants observed in the high-risk group. However, PolyPhen-2 HDIV showed limited utility, underscoring the need to rely on better-calibrated and validated predictors. While low allele frequency is an important filtering criterion, our analysis reinforces that functional and clinical annotations are essential for identifying pathogenic variants.

Together, these findings highlight the utility and limitations of current computational tools and databases in variant interpretation. They also provide a scalable framework for assessing variant burden across cancer susceptibility genes. Continued integration of population genomics, functional validation, and clinical data will be key to resolving uncertain variants and enhancing precision medicine in hereditary breast cancer.

## Data Availability

Data Is publicly available through:-

1. The NYGC 30× GRCh38 1000 Genomes annotated coding and rare-coding flat files were obtained from the IGSR/NYGC 30× data collection (https://www.internationalgenome.org/data-portal/data-collection/30x-grch38).
2. Exome variant data and allele frequencies for breast cancer genes were obtained from gnomAD v4.1.0 (https://gnomad.broadinstitute.org/), which provides publicly accessible, de-identified population genetic data.

## Author contributions

Kanishk Yadav, MS – Conceptualization, Coding, Writing the manuscript, Figures Abhishek Prasad, MD – Editing the manuscript

## Competing interests

The authors have no conflicts of interest to declare.

## Declarations/Ethics

Ethics approval and consent to participate: Not applicable. All data analyzed in this study were obtained from publicly available, de-identified aggregate genomic databases (gnomAD, 1000 Genomes Project, ClinVar). No new data were collected and no identifiable human data, human tissue, or direct human participation were involved.

## Data Availability

All data produced are available online at: https://ftp.1000genomes.ebi.ac.uk/vol1/ftp/data_collections/1000G_2504_high_coverage/working/20201028_3202_raw_GT_with_annot/ and https://gnomad.broadinstitute.org/data

## Notes

### Competing Interest Statement

The authors have declared no competing interest.

### Funding Statement

This study did not receive any funding

### Author Declarations

1)https://ftp.1000genomes.ebi.ac.uk/vol1/ftp/data_collections/1000G_2504_high_coverage/working/20201028_3202_raw_GT_with_annot/ 2)https://gnomad.broadinstitute.org/data

### Summary of Updates

The Figure 3 has been updated.

